# No Laughing White Matter: Cortical Cholinergic Pathways and Cognitive Decline in Parkinson’s Disease

**DOI:** 10.1101/2023.05.01.23289348

**Authors:** Rachel A. Crockett, Kevin B. Wilkins, Sudeep Aditham, Helen M. Brontë-Stewart

**Author notes:** Corresponding author at: Stanford Movement Disorders Center (SMDC), Department of Neurology and Neurological Sciences, 453 Quarry Rd, Stanford University School of Medicine, Stanford, California, USA, 94305; 1. **Funding for Study:** PPMI – a public-private partnership – is funded by the Michael J. Fox Foundation for Parkinson’s Research and funding partners, including, Abbvie, AcureX, Allergan, Amathus, Therapeutics, Asap, Avid, Biogen, Bial Biotech, Biolegend, BlueRock Therapeutics, Bristol-Myers, Calico, Celgene, Cerevel, Coave Therapeutics, Dacapo BrainScience, Jenali Therapeutics, 4D Pharma plc, GE Healthcare, Edmond J. Safra Philanthropic Foundation, Genentech, GlaxoSmithKline, Golub Capital, Gain Therapeutics, Handl Therapeutics, Insitro, Janssen Neuroscience, Lilly, Lundbeck, Merch, MSD Veso Scale Discovery, Neuroscine Biosciences, Pfizer, Piramal Healthcare, Prevail Therapeutics, Roche, Sanofi Genzyme, Servier, Takeda, Teva, UCB, Vanqua Bio, Verily, Voyager Therapeutics, and Yumanity Therapeutics.

## Abstract

**Background:** Approximately one third of recently diagnosed Parkinson’s disease (PD) patients experience cognitive decline. The nucleus basalis of Meynert (NBM) degenerates early in PD and is crucial for cognitive function. The two main NBM white matter pathways include a lateral and medial trajectory. However, research is needed to determine which pathway, if any, are associated with PD-related cognitive decline.

**Methods:** Thirty-seven PD patients with no mild cognitive impairment (MCI) were included in this study. Participants either developed MCI at 1-year follow up (PD MCI-Converters; n=16) or did not (PD no-MCI; n=21). Mean diffusivity (MD) of the medial and lateral NBM tracts were extracted using probabilistic tractography. Between-group differences in MD for each tract was compared using ANCOVA, controlling for age, sex, and disease duration. Control comparisons of the internal capsule MD were also performed. Associations between baseline MD and cognitive outcomes (working memory, psychomotor speed, delayed recall, and visuospatial function) were assessed using linear mixed models.

**Results:** PD MCI-Converters had significantly greater MD of both NBM tracts compared to PD no-MCI (p<.001). No difference was found in the control region (p=.06). Trends were identified between: 1) lateral tract MD, poorer visuospatial performance (p=.05) and working memory decline (p=.04); and 2) medial tract MD and reduced psychomotor speed (p=.03).

**Conclusions:** Reduced integrity of the NBM tracts is evident in PD patients up to one year prior to the development of MCI. Thus, deterioration of the NBM tracts in PD may be an early marker of those at risk of cognitive decline.

## Introduction

The World Health Organization estimates that disability and death due to Parkinson’s disease (PD) is increasing faster than any other neurological disorder (1). With its prevalence doubling in the last 25 years (1), the need for effective treatments are medically and economically vital. The cardinal motor symptoms of PD are bradykinesia, rigidity, and tremor (2). However, among the most disabling non-motor symptoms, cognitive decline is evident in approximately one third of recently diagnosed patients, which increases to up to 50% after five years (3). While medication (4) and deep brain stimulation (5) of the dopaminergic system can provide long-term improvement for PD motor symptoms, treatments for cognitive impairment remain scarce and ineffective.

The nucleus basalis of Meynert (NBM) is at the epicentre of the cortical cholinergic network and is crucial for the cognitive domains most vulnerable in PD, including attention, visuospatial, and executive functions (6). It is known to degenerate early in PD, which was predictive of cognitive but not motor impairment up to five years later (7). Further, Schulz and colleagues (8) identified that greater mean diffusivity, which is indicative of reduced tissue integrity (9), of the NBM region was predictive of PD patients who would go on to develop cognitive impairment up to three years later. Notably, differences in mean diffusivity of other PD affected regions, including the entorhinal cortex, amygdala, hippocampus, insula, and thalamus, were not predictive of cognitive decline. In addition, it is now recognised that some traditionally motor tasks (e.g., gait) require higher-order cognitive control in older adults (10), referred to as the cognitive-motor syndrome. While resistant to dopaminergic treatments, poorer performance on cognitive-motor tasks has also been linked to atrophy of the NBM (11). This indicates that deficits to the cholinergic network may underlie cognitive and cognitive-motor decline in PD.

There are two main pathways emanating from the NBM (see Figure 1). The lateral pathway that travels through the external capsule and uncinate fasciculus to supply the frontal, insula, parietal, and temporal cortices; and a medial pathway, passing through the cingulum to supply the parolfactory, cingulate, pericingulate and retrosplenial cortices (12). Evidence from older adults with vascular cognitive impairment suggests specific associations between cognitive function and the integrity of the NBM tracts but not with NBM volume (13). Nemy and colleagues (14) support these findings by showing the integrity of the lateral and medial pathways had stronger contributions than NBM volume itself to measures of attention and memory in older adults with cerebral small vessel disease, despite their NBM volume being reduced. These findings suggest that integrity of the NBM tracts may be more important than NBM volume for the development of cognitive decline.

**Figure 1.**
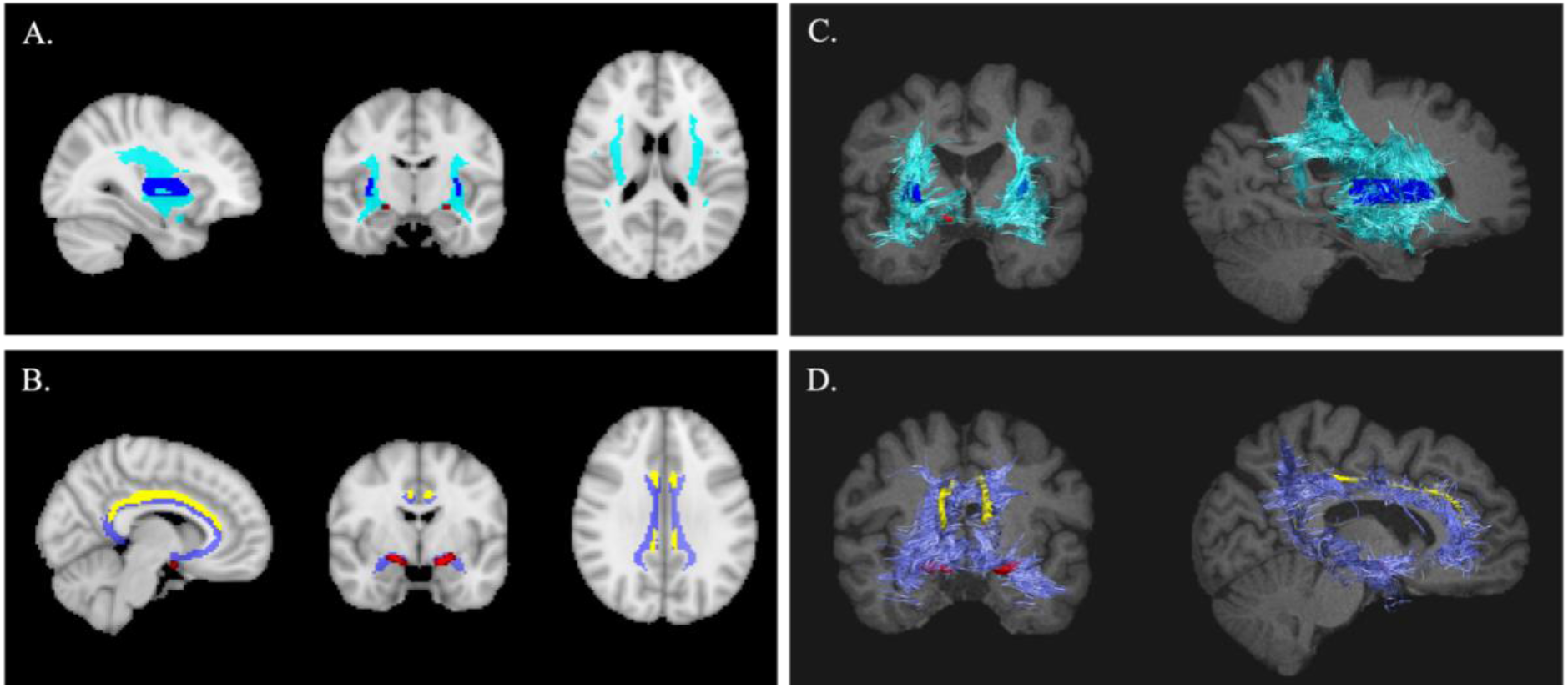
The templates for the lateral (A) and medial (B) tracts in MNI space and an example 3D representation of the lateral (C) and medial (D) template tracts in the subject space. Cyan: lateral tract; Purple: medial tract; Blue: external capsule; Yellow: cingulum; Red: nucleus basalis of Meynert.

Schumacher and colleagues (15) investigated the involvement of these tracts in healthy controls compared to patients with dementia or mild cognitive impairment resulting from Alzheimer’s disease or dementia with Lewy bodies. They demonstrated that while reduced NBM volume was evident in all clinical groups compared to healthy controls, this was less associated with cognitive impairment and was not associated with conversion to dementia. However, they identified reduced integrity of the lateral tract in both the dementia and mild cognitively impaired groups compared to healthy controls, which was associated with impaired global cognition and poorer attention. Importantly, while reduced integrity of the medial tract was not associated with cognitive outcomes, integrity of both tracts was associated with increased risk of progression to dementia. Research is needed to determine whether these differences in the structural integrity of the lateral and/or medial tracts also exist in PD patients who develop cognitive impairment compared to those that remain cognitively healthy. Further, identifying which cognitive domains may be associated with each tract would provide greater insight of the neurobiological mechanisms underlying cognitive and cognitive-motor impairment in PD.

The aim of this study was to investigate the white matter integrity of the lateral and medial NBM tracts in PD patients who develop cognitive impairment compared to those that remain cognitive healthy. In addition, we aimed to determine whether integrity of these tracts may be associated with decline in specific domains of cognition. We hypothesised that: 1) PD patients who develop cognitive impairment will have reduced integrity of the lateral and medial NBM tracts compared to PD patients who remain cognitively intact; and 2) poorer integrity of the lateral tract will be associated with greater decline in memory, visuospatial processing, and executive function, while poorer integrity of the medial tract will be associated with greater decline in memory.

## Methods

### Participants

Thirty-seven participants with PD from the Parkinson’s Progression Markers Initiative (PPMI) database (www.ppmi-info.org/access-data-specimens/download-data) were included in this study. Participants were included in the PPMI study if they: 1) were ≥30 years old; 2) had PD for at least 2 years prior to screening; 3) were not currently on or expected to start PD medication for at least 6 months after baseline testing; 4) were not currently or planning to become pregnant during the study; and 5) were able to provide informed consent. They were excluded if they: 1) were currently taking PD medication; 2) had other atypical PD syndromes (e.g., metabolic disorders, encephalitis, or other neurodegenerative disease); 3) were clinically diagnosed with dementia; or 4) had any other medical or psychiatric condition which might preclude participation. Additional inclusion criteria for the current study required participants to have: 1) undergone magnetic resonance imaging with both a structural T1-weighted and diffusion weighted scan; 2) no cognitive impairment at baseline indicated by a score >26 on the Montreal Cognitive Assessment (MoCA); and 3) completed cognitive testing at one year follow up.

### Measurements

#### Montreal Cognitive Assessment (MoCA)

The MoCA (16) is a measure of global cognition and has a high level of sensitivity and specificity for MCI (score <26/30). It involves tests of eight cognitive domains: 1) attention; 2) executive functions; 3) memory; 4) language; 5) visuo-constructional skills; 6) conceptual thinking; 7) calculations; and 8) orientation. It is scored out of 30 with lower scores indicative of poorer cognitive function. Education level is accounted for in the scoring.

Using the MoCA scores at one year follow up, the PD participants were further characterized as having developed mild cognitive impairment (MCI-converters) or having maintained cognitive health (PD no-MCI).

#### Working Memory

The letter number sequencing sub-component of the Weschler Adult Intelligence Scale (WAIS) (17) was used to assess working memory. Participants hear a random sequence of numbers and letters and are asked to recall first the numbers in ascending order, followed by the letters in alphabetical order. A point is scored for every correct recall and the test is stopped after three failed attempts at recalling a sequence. There is a maximum score of 21 with lower scores indicative of poorer working memory.

#### Processing and Psychomotor Speed

The symbol digit modalities test (18) was used to assess psychomotor and processing speed. Participants were presented with abstract symbols corresponding to the numbers 1-9. They are then asked to write or orally report the correct number that corresponds to a series of pre-defined symbols. They are scored based on the number of correct symbols drawn in 90 seconds with lower scores indicative of poorer information processing, and psychomotor speed.

#### Delayed Recall

Delayed recall was assessed using the Hopkins Verbal Learning Test – Revised (19). Participants are read aloud 12 nouns and asked to immediately recall as many of the words as they can remember. This is repeated three times with the same words each time. After a delay of 20-25 minutes participants are asked to name as many words as they can remember. They are not informed about the delay prior to the trial. Lower scores indicate poorer delayed recall.

#### Visuospatial Processing

The Benton judgement of line orientation (20) was used to assess visuospatial processing. Participants are shown pairs of lines at different angles with sections omitted and asked to identify the matching line orientations from a set of 11 numbered radii that form a semi-circle. Both lines must be matched correctly for that pair to be scored correctly. There is a maximum raw score of 15 with higher scores indicative of better visuospatial processing.

For all cognitive measures, the derived scores were used whereby the raw scores were adjusted for age based on normative data. Change scores for all cognitive outcomes were calculated by subtracting baseline score from those at one year follow up.

#### Magnetic Resonance Imaging (MRI) Acquisition

The MRI scans were acquired using standardized procedures across sites on a Siemens 3T scanner. The 3D T1-weighted (T1-w) images consisted of 192 slices, with a 1mm3 voxel size, an acquisition matrix of 256 × 256, a repetition time (TR) of 2300ms, and an echo time (TE) of 2.98ms. The 2D diffusion-weighted echo planar images (DWI) consisted of ∼80 slices, with a of 2mm3 voxel size. They had a TR of ∼10000ms, TE of ∼80ms, flip angle of 90°, acquisition matrix of 128 × 128, *b* = 1000 /mm2 with one *b*0 image, and 64 gradient directions.

#### Tractography Pipeline

Preprocessing of the MRI data was completed using the FMRIB Software Library (FSL) (21). The tractography pipeline was designed based on methods previously shown to be successful for segmenting the two NBM tracts of interest in older adults with and without neurodegenerative disease (14,15).

Removal of non-brain structures was completed using HD-BET (22) on the T1-w, DWI and the *b*0 slice. All images were then converted to the anterior commissure-posterior commissure (ACPC) line using the rigid linear conversion from T1-w to the MNI ACPC line with six degrees of freedom as the reference transformation (23). The gradient direction matrices (bvecs) were also rotated to account for this transformation.

The brainstem was extracted using FSL’s First automated segmentation tool (24), and the hemisphere mask was hand drawn in FSLeyes by identifying the midline of the brain in the coronal plane on the subject scans. All other regions were identified using predetermined atlases in MNI space, which were then transformed to subject space. The internal capsule, external capsule, and cingulum were extracted from the Johns Hopkins University white matter atlas (25), the anterior commissure from FSL’s XTRACT atlas (26), and the NBMs from the Ch4 basal forebrain regions identified by stereotaxic probabilistic maps from 10 high quality post-mortem brains (27). After preprocessing the data with bedpostX, probabilistic tractography was completed using the standard 5000 samples and 0.2 curvature threshold settings in probtrackX with NBM as the seed region, and either the ipsilateral external capsule (lateral tract) or cingulum (medial tract) as the waypoint regions of inclusion. The anterior commissure, internal capsule, brain stem, and hemisphere were specified as regions of avoidance.

All extracted tracts were converted to MNI space using nonlinear transformation. The tracts for each hemisphere were flipped and overlayed onto the contralateral hemisphere to create consistent templates across hemispheres. A template of both the lateral and medial tracts was created for each hemisphere that contained tracts present in at least 50% of the participants (see Figure 1.A and 1.B). These template tracts were then transformed back into the subject space and binarized to create participant specific tract masks (see Figure 1.C and 1.D). A map of the mean diffusivity (MD) of each voxel was created using DTIFit and the template tracts overlayed to determine the average MD of each tract for every participant.

To determine if differences in tract integrity may be reflective of brain-wide neurodegeneration, the MD of the internal capsule was also used as a control comparison to identify if there were any differences between groups in a region unrelated to the tracts of interest.

#### Statistical Analysis

All statistical analyses were completed in R version 4.2.2. Two analyses of covariance (ANCOVA) models were used to determine between group differences in the MD of the lateral and medial tracts. Age, disease duration, and sex were included as covariates and hemisphere was specified as a random variable. Linear mixed models were used to determine whether the MD of either tract was associated with baseline and/or changes in cognitive function at one year follow up. Age was already controlled for in the cognitive outcomes and sex, disease duration, and baseline cognitive performance (change scores only) were included as covariates with hemisphere included as the random variable. A false discovery rate correction (q=.05) was used to control for multiple comparisons.

## Results

The total sample (N=37) included nine females with a mean (SD) age of 60.1 (9.9) years. There were 21 participants characterized as PD no-MCI, and 16 MCI-converters (see Table 1). On average the PD no-MCI group had a 0.1 (1.4) decline in MoCA score from baseline to one year follow up, while the PD MCI-Converters showed a 5.3 (3.3) decline. One extreme outlier (Mean ± 3SD) was detected for the MD of the lateral tract in the PD no-MCI group. However, removal of this outlier did not change the significance of the results and thus was kept in the final analyses. No outliers were detected in either the baseline or change scores for the cognitive outcomes. The MCI-Converters were significantly older than the PD no-MCI group (t= -7.2, p<0.001) and were more likely to be males (χ^2^= 4.3, p=0.04). Both age and sex were controlled for in the analyses.

**Table 1.** Participant Characteristics

|  |  | Mean (SD) or n(%) |  |
| --- | --- | --- | --- |
|  |  | PD no-MCI | MCI-Converters |
| n |  | 21 | 16 |
| Age (yrs) |  | 54.8 (9.2) | 67.1 (5.3) |
| Sex (Female) |  | 7 (33.3) | 2 (12.5) |
| Education (yrs) |  | 16.0 (9.2) | 14.9 (3.3) |
| MoCA (/30) |  | 28.9 (1.3) | 27.6 (1.2) |
| Symbol Digit Modalities Test | Baseline | 48.1 (9.2) | 33.1 (9.8) |
|  | Follow Up | 47.8 (9.6) | 33.6 (11.4) |
|  | Change | -.29 (6.3) | .50 (4.7) |
| Letter Number Sequencing | Baseline | 11.3 (2.8) | 9.6 (2.5) |
|  | Follow Up | 11.0 (2.6) | 8.8 (2.8) |
|  | Change | -.33 (2.0) | -.81 (3.4) |
| HVLT - Delayed Recall | Baseline | 9.2 (2.5) | 6.8 (3.3) |
|  | Follow Up | 9.2 (2.4) | 6.4 (2.7) |
|  | Change | -.05 (2.8) | -.44 (2.4) |
| Benton Judgement of Line Orientation | Baseline | 27.4 (3.1) | 23.0 (6.2) |
|  | Follow Up | 27.0 (3.7) | 24.4 (6.0) |
|  | Change | -.38 (3.9) | 1.4 (4.0) |

### Between-Group Comparisons of the Lateral and Medial Tracts

There was a significant effect of group on the MD of both the lateral (F=35.3, p<0.001) and medial (F=34.7, p<0.001) tracts (see Figure 2). Whereby, the PD MCI-Converters had greater MD of both tracts compared to the PD no-MCI patients. Results of the control analyses showed no significant difference between PD groups in the MD of the internal capsule (F=3.8, p=0.06), suggesting that the above results were tract-specific and not due to brain-wide atrophy as a result of PD.

**Figure 2.**
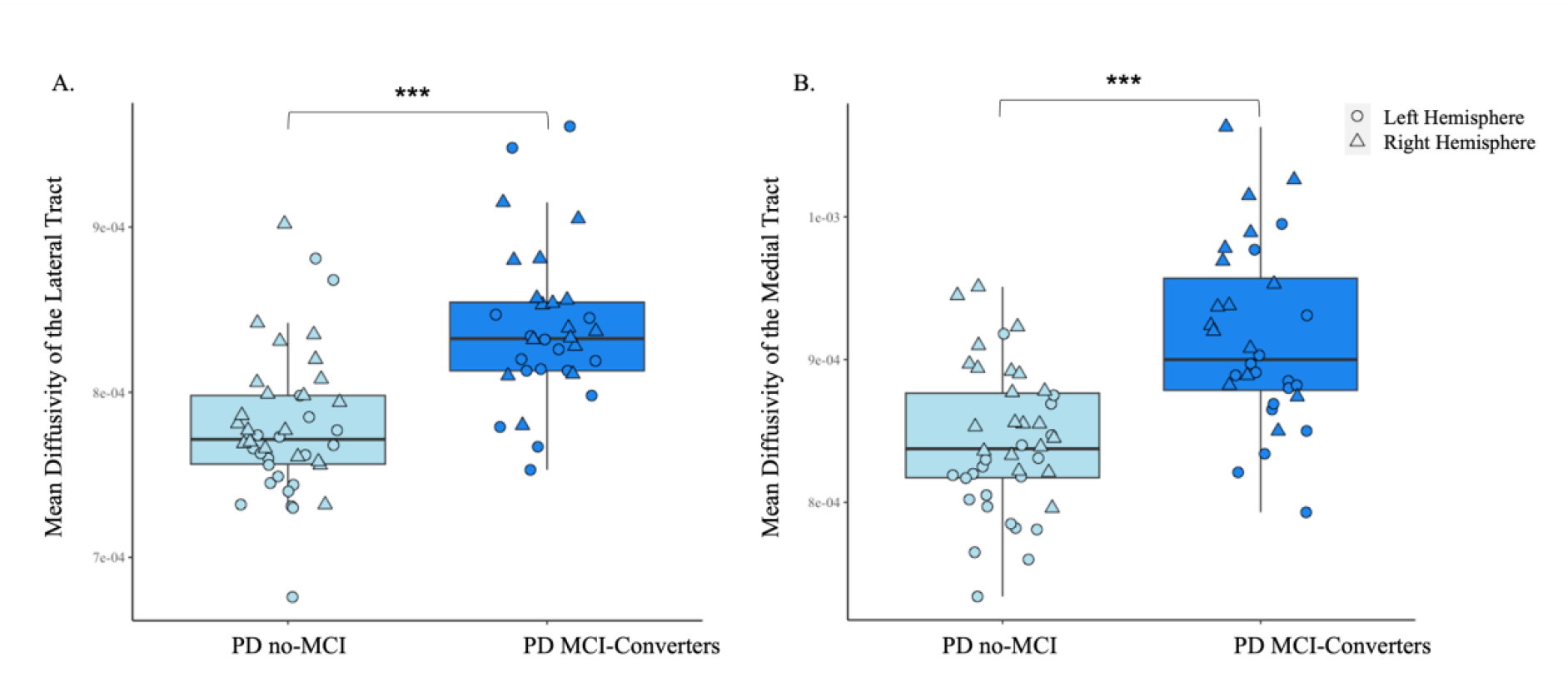
Comparison of the mean diffusivity of the lateral (A) and medial (B) tracts of the nucleus basalis of Meynert (NBM) for Parkinson’s disease with no mild cognitive impairment (PD no-MCI) and Parkinson’s disease patients who developed mild cognitive impairment at one year follow up (PD MCI-Converters). Circle = Left NBM tract; Triangle = Right NBM tract. ***p<0.001

### Associations between MD of the NBM Tracts and Cognitive Outcomes

Baseline performance on the Benton judgement of line orientation test was associated with MD of the lateral tract (t= -2.0, p=0.05) while performance on the symbol-digit modalities test was associated with MD of the medial tract (t= -2.2, p=0.03). No other cognitive measures at baseline were associated with MD of either tract (see Table 2). In addition, greater MD of the lateral tract was associated with greater decline on the letter-number sequencing task (t= -2.1, p=0.04). There were no other associations identified between change in cognitive outcomes and MD of either tract (see Table 2).

**Table 2.** Linear mixed models controlling for sex, disease duration, and baseline cognitive outcome (for change scores), in age-adjusted cognitive outcomes.

|  |  | Lateral Tract | Medial Tract |
| --- | --- | --- | --- |
| **Baseline** |  |  |  |
| Symbol Digit Modalities Test | t | -1.8 | -2.2 |
|  | p | .07 | .03* |
| Letter Number Sequencing | t | .28 | -.18 |
|  | p | .78 | .86 |
| HVLT – Delayed Recall | t | -.24 | -.30 |
|  | p | .81 | .76 |
| Benton Judgment of Line Orientation | t | -2.0 | -1.9 |
|  | p | .05* | .07 |
| **Change** |  |  |  |
| Symbol Digit Modalities Test | t | .02 | .15 |
|  | p | .98 | .88 |
| Letter Number Sequencing | t | -2.1 | -1.7 |
|  | p | .04* | .09 |
| HVLT – Delayed Recall | t | -.03 | -1.1 |
|  | p | .98 | .27 |
| Benton Judgment of Line Orientation | t | 1.1 | .99 |
|  | p | .27 | .33 |

\*p≤0.05

These associations did not survive corrections for multiple comparisons and thus are considered trends rather than significant findings.

## Discussion

This study is the first to investigate the differences in integrity of the lateral and medial NBM tracts in patients with PD who develop MCI compared to those that remain cognitively healthy. We identified that PD patients who developed MCI had greater MD of both the lateral and medial tracts than those who did not develop MCI. We also identified trends between the MD of the lateral and medial tract with poorer visuospatial processing and psychomotor speed respectively. An additional trend was also identified between higher MD of the lateral tract and greater decline in working memory.

Our finding that the lateral and medial tracts are more intact in the PD no-MCI groups compared to PD patients who developed MCI is consistent with the findings from the aging and dementia literature (14,15). It is also notable that these differences were observed prior to the development of MCI, providing further evidence that degeneration of the cholinergic network is an early predictor of cognitive decline in PD (7,8). Specifically, we identified trends in the association between greater MD of the lateral tract with poorer visuospatial function at baseline, and with greater decline in working memory at one year follow-up. Lesioning the NBM in non-human primates resulted in significant impairments in visuospatial performance (28). Our findings extend the role of this region in visuospatial function by suggesting that this may be specific to the lateral tract. Considered an executive function task, working memory requires significant involvement of the prefrontal cortex (29), which is supplied by the lateral NBM tract (6). Previous research has also highlighted the involvement of these tracts in attention (30). While we were not able to assess this relationship with a specific attention task, it has been widely accepted that performance of working memory requires significant attentional resources (31). Thus, it is likely that reduced integrity of the lateral tract impacts the cholinergic supply to brain regions needed for optimal executive function and attention.

Our results are somewhat consistent with previous research that found no association between the integrity of the medial tract and cognitive function in neurodegenerative disease despite there being involvement of this tract in cognitive status (15). However, we did identify a trend between greater MD of the medial tract and poorer psychomotor speed. Performance on a similar psychomotor task was associated with greater activation of areas of the prefrontal, parietal and cingulate cortices in older adults (32). All areas that are supplied by the medial NBM tract. It is important to note that the relationships discussed between tract integrity and cognitive function were not significant after controlling for multiple comparisons. Consistently, findings in both older adults and neurodegenerative populations have demonstrated associations between the integrity of the NBM tracts and performance on measures of global cognition (7,8,14,15,30). Therefore, it is possible that degeneration of these tracts may underlie a broader decline in cognition that is less consistently detectable by specific cognitive domain measures.

We found no association between either tract and delayed recall. This is inconsistent with previous research that identified associations between the lateral tract and NBM volume with delayed recall (7,14). However, this may be due to differences in the clinical status of the sample (healthy older adults with cerebral small vessel disease) and/or differences between associations of the NBM volume itself compared to the specific integrity of the tracts. The PD patients in this study were de-novo. Thus, while there may be early evidence of differences between those who do and do not develop MCI, the contribution of these tracts to specific cognitive functions may become more apparent at later disease stages. Further research is needed to elucidate the exact role of this tract in PD-related cognitive decline.

This study is not without limitations. Firstly, we were unable to investigate differences in tract integrity more than one year before potential MCI conversion. Therefore, it is possible that these differences may be evident even earlier than suggested by our findings. In addition, the patients in this study were at early stages of PD. While this does provide evidence of cholinergic decline at very early disease stages, the differences seen between PD patients with and without development of MCI cannot be extended to more severe disease progression. Finally, due to study designs beyond our control, we were unable to investigate a broader range of cognitive functions that may have provided greater understanding of the domains affected by degeneration of the lateral and medial NBM tracts specifically.

## Conclusion

PD is predominantly a movement disorder caused by neurodegeneration of the dopaminergic network. However, a large percentage of these patients will also develop cognitive decline which may progress to dementia. Our findings suggest that this decline in cognition may be due to additional degeneration of the cholinergic network. Notably, these deficits in cholinergic white matter tract integrity were evident up to one year prior to the development of MCI. Thus, suggesting that greater MD of the NBM tracts may be an early indicator of those at risk of cognitive decline.

## Data Availability

Data used in the preparation of this article were obtained from the Parkinson's Progression Markers Initiative (PPMI) database (www.ppmi-info.org/access-data-specimens/download-data). For up-to-date information on the study, visit www.ppmi-info.org.

## Authors’ Roles

RAC, KBW, and HMBS were involved in the design, data analyses, and interpretation of results. SA was involved in the data analyses. RAC wrote the first draft of the manuscript.

KBW, SA and HMBS wrote portions of the manuscript and all authors critically reviewed and approved the manuscript.

## Funding

The authors are funded by two National Institutes of Health (NIH): National Institute of Neurological Disorders and Stroke (NINDS) research grants (RAC, KBW and HMBS: UG3NS128150; SA and HMBS: UH3NS107709).

## Notes

**Conflict of Interest:** The authors have no conflicts of interest to disclose.

### Competing Interest Statement

The authors have declared no competing interest.

